# An effect assessment of Airborne particulate matter pollution on COVID-19: A multi-city Study in China

**DOI:** 10.1101/2020.04.09.20060137

**Authors:** Bo Wang, Jiangtao Liu, Shihua Fu, Xiaocheng Xu, Lanyu Li, Yueling Ma, Ji Zhou, Jinxi Yaoc, Xingrong Liu, Xiuxia Zhang, Xiaotao He, Jun Yan, Yanjun Shi, Xiaowei Ren, Jingping Niu, Bin Luo, Kai zhang

## Abstract

**Objective:** Coronavirus disease 2019 (COVID-19) is a serious infectious disease, which has caused great number of deaths and health problems worldwide. This study aims to examine the effects of airborne particulate matter (PM) pollution on COVID-19 across China.

**Methods:** In this study, we obtained confirmed cases of COVID-19, the data of airborne ambient PM with aerodynamic diameter ≤ 2.5 μm (PM_2.5_) and ≤ 10 μm (PM_10_), ambient temperature (AT), absolute humidity (AH) and migration scale index (MSI) in 72 cities of China (excluded Wuhan city) on a daily basis, each of which confirmed more than 50 cases from January 20th to March 2nd, 2020. We applied a two-stage analysis. Generalized additive models with quasi-Poisson distribution was first fitted to estimate city-specific effects of PM_10_ and PM_2.5_ on daily confirmed COVID-19 cases while controlling AT, AH and MSI. Then, we used meta-analysis to generate the pooled effect estimates from city-specific results.

**Results:** During the study period, there were a total of 24 939 COVID-19 cases, most of which were reported in Hubei Province. In our meta-analysis, we found each 10 μg/m^3^ increase in concentration of PM_2.5_ and PM_10_ in single day lag (from lag 0 to lag 7 and lag 14) were positively associated with confirmed cases of COVID-19, not including PM_10_ at lag 5, lag 6 and lag 7, and PM_2.5_ at lag 5, lag 6. Similar trend was also found in different cumulative lag days (from lag 01 to lag 07 and lag 014). The effects of PM_2.5_ and PM_10_ on daily COVID-19 confirmed cases are statistically significant for three cumulative lag periods over 3, 7 and 14 days with the greatest effect over 14 days. The estimated RRs of which were 1.64 (95% CIs: 1.47, 1.82) and 1.47 (95% CIs: 1.34, 1.61) with each 10 μg/m^3^ increase in concentrations of PM_2.5_ and PM_10_, respectively. In addition, we found that the effects of PM_2.5_ on daily confirmed cases were greater than PM_10_ in all included lag days.

**Conclusions:** This nationwide study suggests that airborne PM pollution likely increases the risk of getting COVID-19 in China.

## 1 Introduction

As of March 27, 509 164 cases of coronavirus disease 2019 (COVID-19) have been confirmed globally with a case fatality rate of 4.58%(WHO 2020), indicating its higher infectivity and lethality. COVID-19 is caused by the 2019 novel coronavirus (SARS-CoV-2), the characteristics of which are very similar to influenza viruses and severe acute respiratory syndrome-associated coronavirus (SARS-CoV). Like influenza and severe acute respiratory syndromes (SARS), environmental factors play a role for their spread and transmission (Chen et al. 2017; Wong et al. 2010). It is confirmed that influenza is a seasonal-depended respiratory disease, causing great health problems among human every year, particularly during winter and early spring (Huang et al. 2017; Wang et al. 2020). COVID-19 was also first confirmed in winter with low temperature and low humidity, which along with small diurnal temperature range were assessed to be the most important environmental factors favoring its transmission (Wang et al. 2020). Dry and cold environment favors SARS-Cov-2 to survive and transmit in droplet or in the form of aerosol (Araujo and Naimi 2020).

Previous studies show that aerosol particles emitting from coughing by influenza patients contain high level of influenza virus and these particles are within the respirable size range (Lindsley et al. 2010). These aerosols having virus are easy to transmit among individuals. It is reported that aerosols from highly virulent pathogens like severe acute respiratory syndrome-coronavirus (SARS-CoV) could travel more than six feet(Kutter et al. 2018). Also, these aerosols are likely composed of airborne pollution particles and attached virus droplets, which promote the spread of pathogen like influenza viruses (Mori et al. 2017). An ecologic analysis found that there were positive relationship between PM_2.5_ concentration and influenza-like-illness risk in Beijing (Feng et al. 2016). In particular, the concentrations of airborne ambient particulate matter (PM) with aerodynamic diameter ≤ 2.5 μm (PM_2.5_) were reported to be significantly associated with daily human influenza cases (Liang et al. 2014; Wong et al. 2010) and respiratory syncytial virus infection (Rich et al.; Vandini et al. 2013). In addition to influenza, the SARS break in 2003 was also found related to air pollution, and the levels of PM with aerodynamic diameter ≤ 10 μm (PM_10_) were positively associated with the SARS mortality (Kan et al. 2005). Like SARS-CoV and influenza viruses, a recent study reported that SARS-CoV-2 was detectable in aerosols for up to three hours, including both liquid and solid aerosol (van Doremalen et al. 2020). Therefore, COVID-19 transmission is likely affected by airborne airborne PM.

Environmental factors were related to the incidence of COVID-19 and SARS, and previous study reported that the population migration was significantly associated with COVID-19 confirmed cases(ZL et al. 2020). Thecontrolling of population migration should be critical to study the association between COVID-19 and environmental factors(Chen et al. 2020). Concerning about that, we collected population migration scale index (MSI) to represent the population migration level and controlled it in the fitted general linear model with binomial negative distribution to examine the associations between air airborne PM pollution and COVID-19 confirmed cases.

## 2 Methods

### Data collection

Using R package “nCov2019” (Wu et al. 2020), we obtained the daily COVID-19 confirmed cases of 72 cities in China, each of which confirmed more than 50 cases from January 20th to March 2nd, 2020. The data of ambient airborne PM, including PM_10_ and PM_2.5_ were obtained from Data Center of Ministry of Ecology and Environment of the People’s Republic of China. At the same time, the data of ambient temperature (AT) and relative humidity were collected from Shanghai Meteorological Bureau. Considering the critical role of population migration in the transmission of the coronavirus, MSI of each city was collected from Baidu migration map (https://qianxi.baidu.com/) which was calculated to represent the scale of population migration. Besides, absolute humidity (AH) was controlled in the models, and it was calculated via the methods reported previously(Tao et al. 2018; Davis et al. 2016).

### Statistical Analysis

We conducted a two-stage analysis to estimate the effects of airborne PM on COVID-19 cases.

In the first stage, we fitted generalized additive models with quasi-Poisson distribution to estimate the associations between airborne PM pollution and daily counts of confirmed case in each city by controlling daily average AT, AH and MSI. The models were fitted based on R software (version 3.6.0) and the “mgcv” package (version 1.8-31). The model framework was as follows:

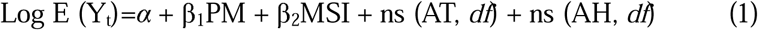

In this model, *t* refers to the day of the observation; Y_t_ is the observed daily confirmed case counts on day *t*; E(Y_t_) is the expected daily confirmed case counts on day t; α is the intercept; β represents the regression coefficient; airborne PM, including PM_10_ and PM_2.5_, represents concentrations at day *t*. We controlled 3-day moving average MSI in the models and used natural smooth functions with 6 *df* for 3-day moving average AT and 3 *df* for 3-day moving average AH to control potential nonlinear and lagged confounding effects of weather conditions. Because the latent period of COVID-19 ranges from 1 to 14 days, mostly 3 to 7 days (Lin and Li 2020), we chose to estimate the single day lag effects (from lag 0 to lag 7 and lag 14) and cumulative lag effects (from lag 01 to lag 07 and lag 014). The results were expressed as the relative risk (RR) and 95% confidence intervals (CIs) for the per 10 μg/m^3^ increase in PM_2.5_ and PM_10_ concentrations.

In the second stage, we used random effects meta-analysis to combine all city-specific results. The subgroup analysis was conducted in different scenarios. First, we explored the effects in different case counts groups. Second, we divided the cities into different groups according to the thresholds of particulate matter levels (PM_10_, 50 μg/m^3^; PM_2.5_, 35 μg/m^3^) of the National Ambient Air Quality Standards in China (GB 3095-2012)(China 2016) and estimated the effects respectively. Third, due to the stricter control measures in Hubei than other cities, we separately analyzed the effects in the cities from Hubei province and the cities outside Hubei.

The meta-analysis was performed based on STATA software (version 11, StataCorp LLC, USA). All statistical tests were two-sided and P-values with less than 0.05 were considered statistically significant.

## 3 Results

In these 72 cities, 43 cities confirmed more than 100 cases and 29 cities confirmed less than 100 cases, among which the 12 Hubei cities (not include Wuhan) confirmed the most cases (16759, 67.20%). The daily average concentration of PM_10_ and PM_2.5_ showed a similar trend, both declined significantly after January 23 (Fig.1).

**Fig. 1.**
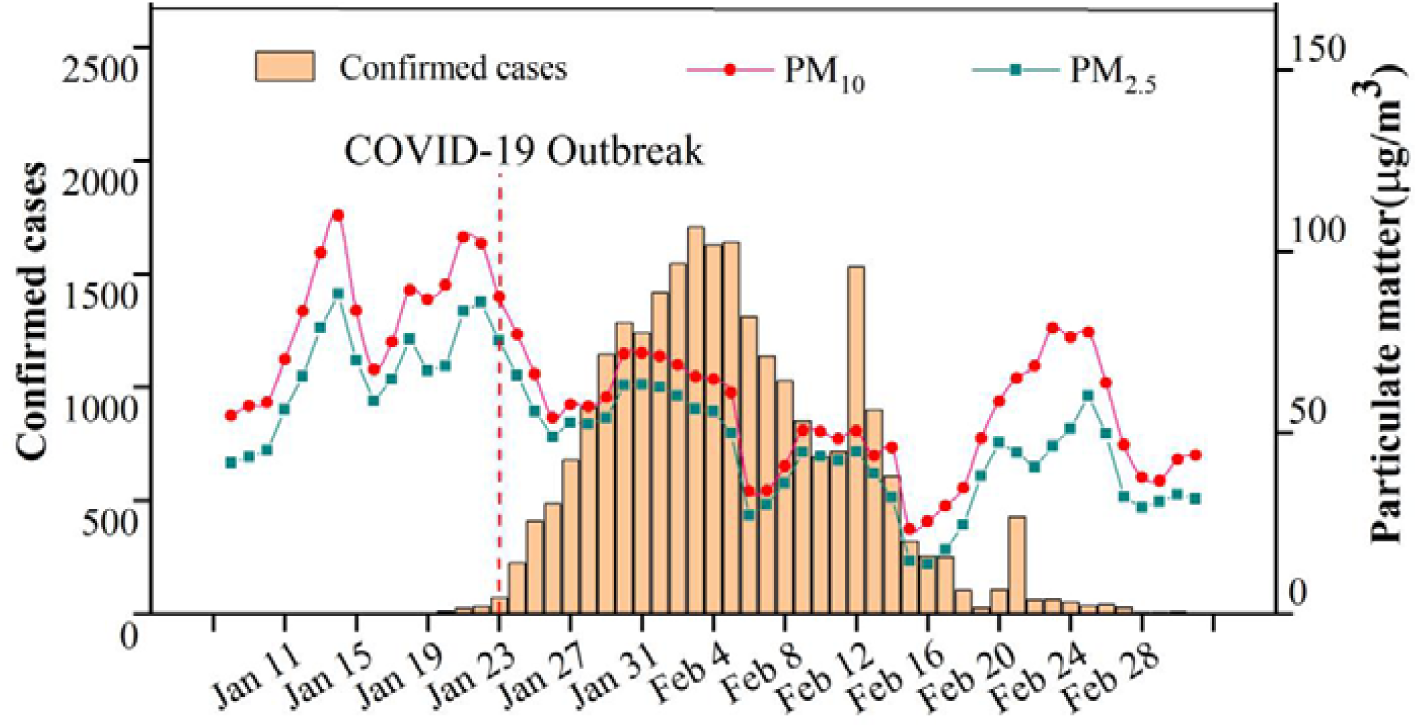
Trends of daily particulate matter levels and daily confirmed COVID-19 cases in 72 Chinese cities during January 08 to March 02 2020.

As shown in Fig.2, we used meta-analysis to pool all the city-specific effects together according to the stratification via total case counts more or less than 100 cases. There were significantly positive associations between the daily confirmed COVID-19 cases and PM_10_, PM_2.5_. For PM_10_ and PM_2.5_, the single-day overall effects were the strongest at lag 3, the corresponding RRs of which were 1.08(95% CIs: 1.01, 1.11) and 1.11 (95% CIs: 1.08, 1.14), respectively. In cumulative lag effects, the pooled estimates of 72 cities were all significant and the strongest effects for both PM_10_ and PM_2.5_ appeared in lag 014 and the RRs of each 10 μg/m^3^ increase were 1.47(95% CIs:1.34, 1.61) and 1.64 (95% CIs:1.47, 1.82), respectively. Those cities with more than 100 confirmed cases showed higher risk of COVID-19 related to PM_10_ and PM_2.5_ compared with cities with less than 100 cases. Additionally, the overall effect is much higher for PM_2.5_ than the PM_10_.

**Fig. 2.**
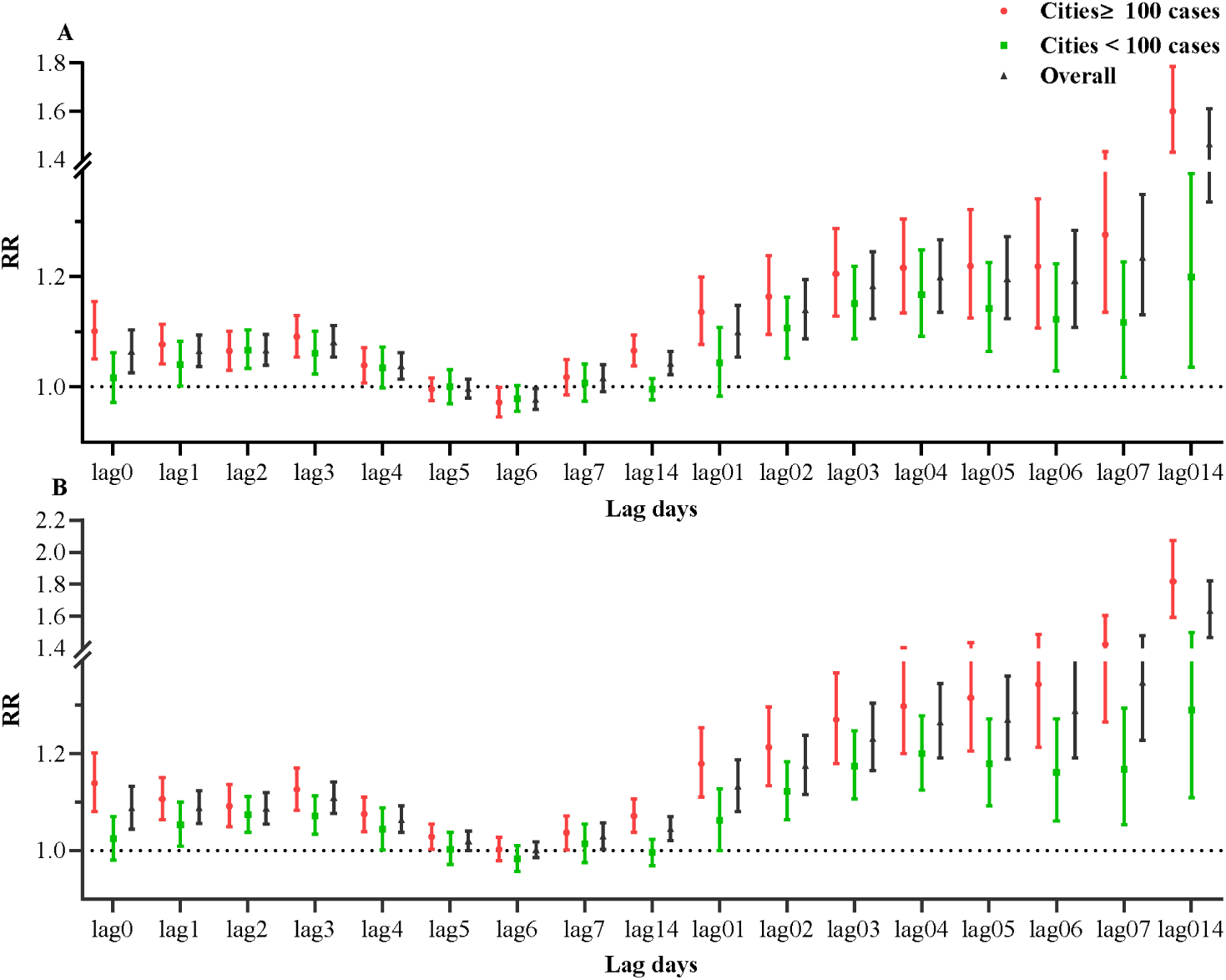
Associations between airborne PM concentration and confirmed cases stratified by total case counts in 72 cities of China during January 20 to March 02 2020. Note: (A) PM_10_; (B) PM_2.5_. The results were expressed as the relative risk (RR) and 95% confidence intervals (CIs) for the per 10 μg/m^3^ increase in PM_2.5_ and PM_10_ concentrations.

Fig.3 showed the associations between airborne PM pollution and confirmed cases stratified by airborne PM concentrations in 72 cities of China. For PM_2.5_, there were 49 cities with an average PM_2.5_ concentration greater than 35 μg/m^3^ and 23 cities less than 35 μg/m^3^. For PM_10_, there are 37 cities with an average PM_10_ concentration greater than 50 μg/m^3^ and 35 cities with the concentrations smaller than 50 μg/m^3^. Also, significant positive effects were found both in the PM_2.5_ and PM_10_ at different levels, but stronger effects were found in cities with the concentration of PM_2.5_ and PM_10_ under national standards.

**Fig. 3.**
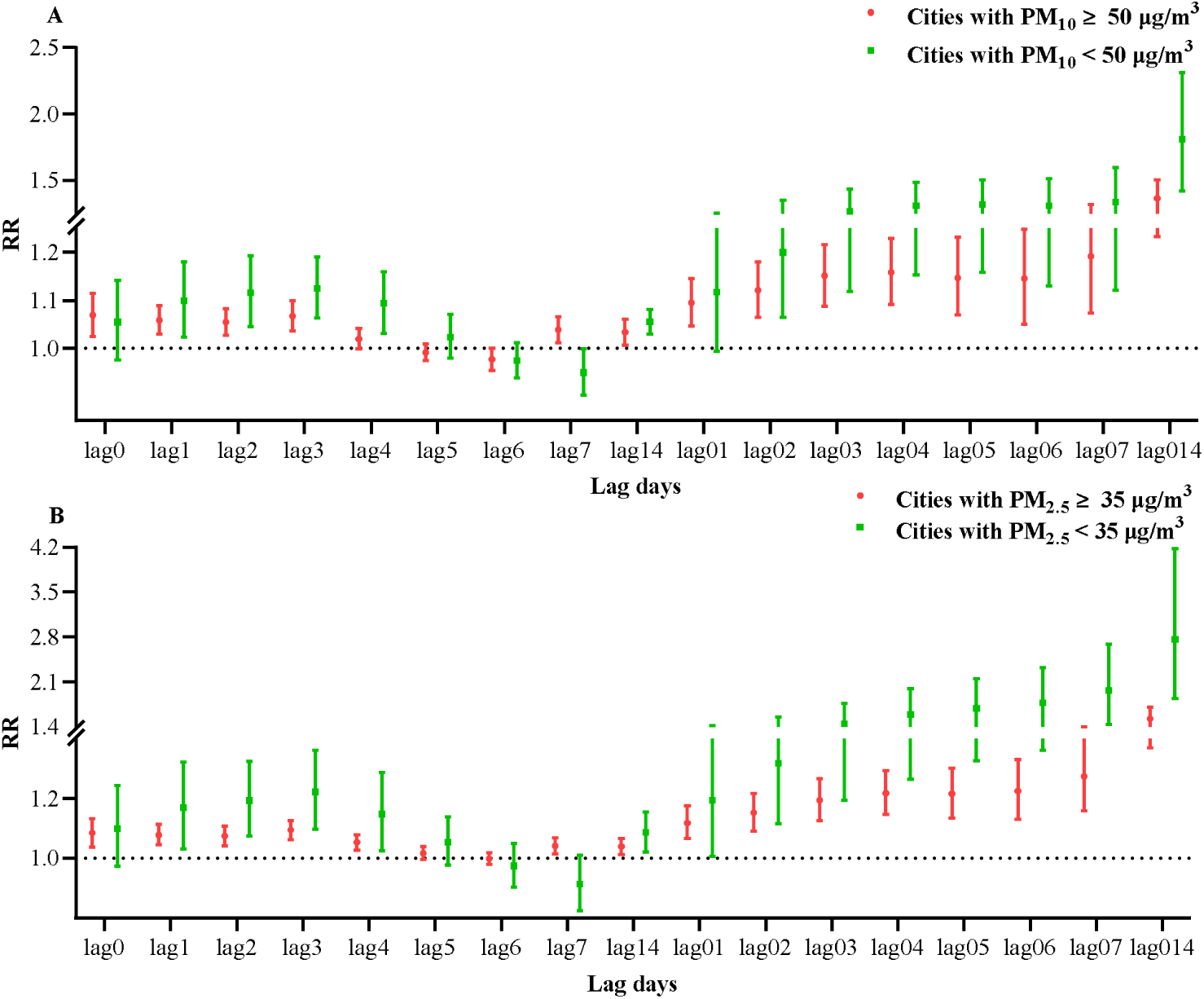
Associations between airborne PM pollution and confirmed cases stratified by airborne particulate matters concentrations in 72 cities of China during January 20 to March 02 2020. Note: (A) PM_10_; (B) PM_2.5_. The results were expressed as the relative risk (RR) and 95% confidence intervals (CIs) for the per 10 μg/m^3^ increase in PM_2.5_ and PM_10_ concentrations.

**Fig. 4.**
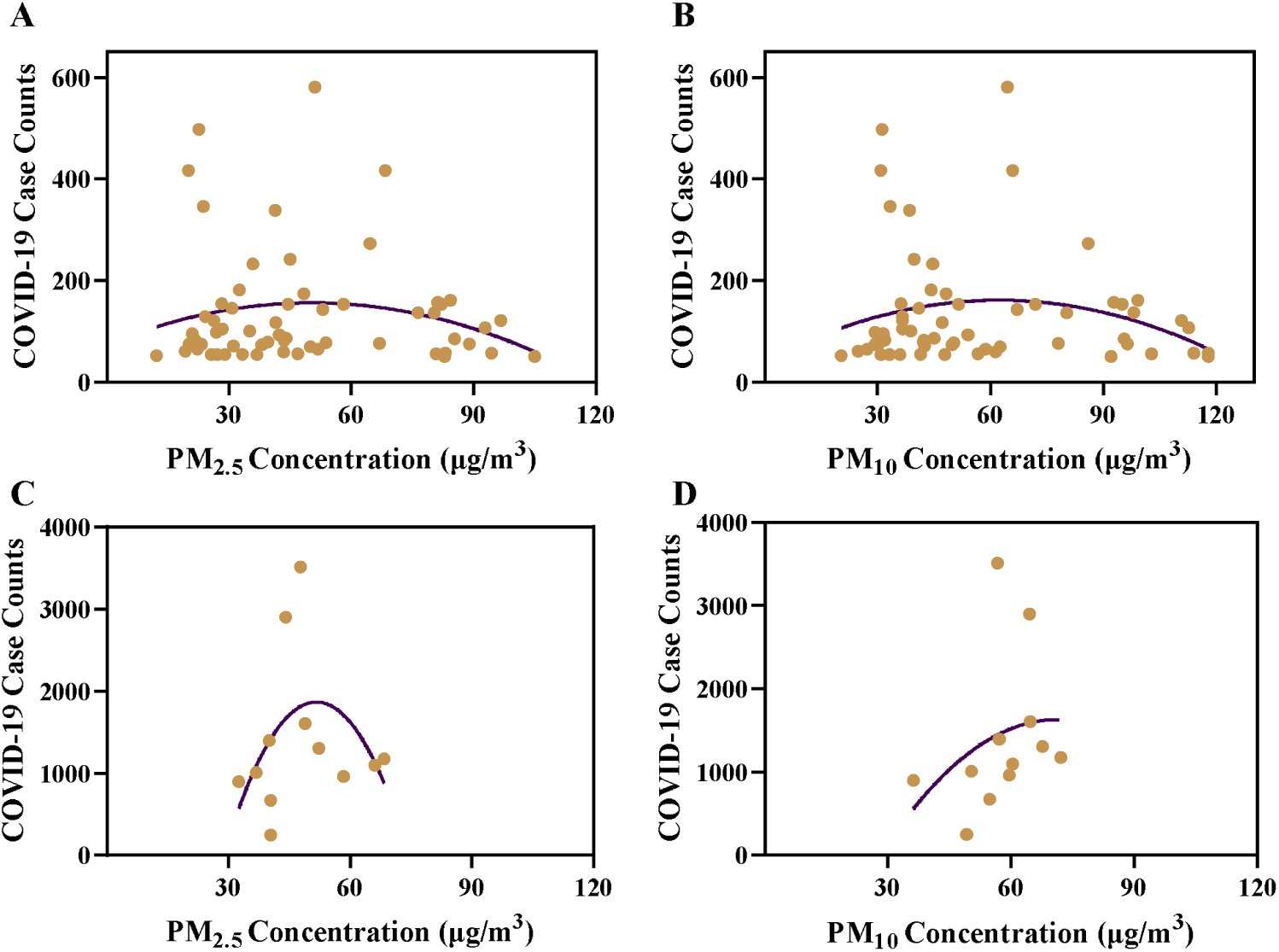
Associations between COVID-19 confirmed case counts and airborne PM concentrations in 72 cities of China during January 20 to March 02 2020 Note: (A) 60 cities outside Hubei Province, PM_2.5_, Curve formula: Y=73.99+3.249_*_X-0.03226_*_X^2^, R^2^= 0.02917; (B) 60 cities outside Hubei Province, PM_10_, Curve formula: Y=40.35+3.875_*_X-0.03109_*_X^2^, R^2^= 0.04113; (3) 12 cities in Hubei Province, PM_2.5_, Y=-7617+366.9_*_X-3.548_*_X^2^, R^2^=0.2006; (4) 12 cities in Hubei Province, PM_10_, Y=-2828+125.2_*_X-0.8804_*_X^2^, R^2^=0.1038. In each figure, the purple line represents second order polynomial curves and the brown dots represent cities.

**Fig.5.**
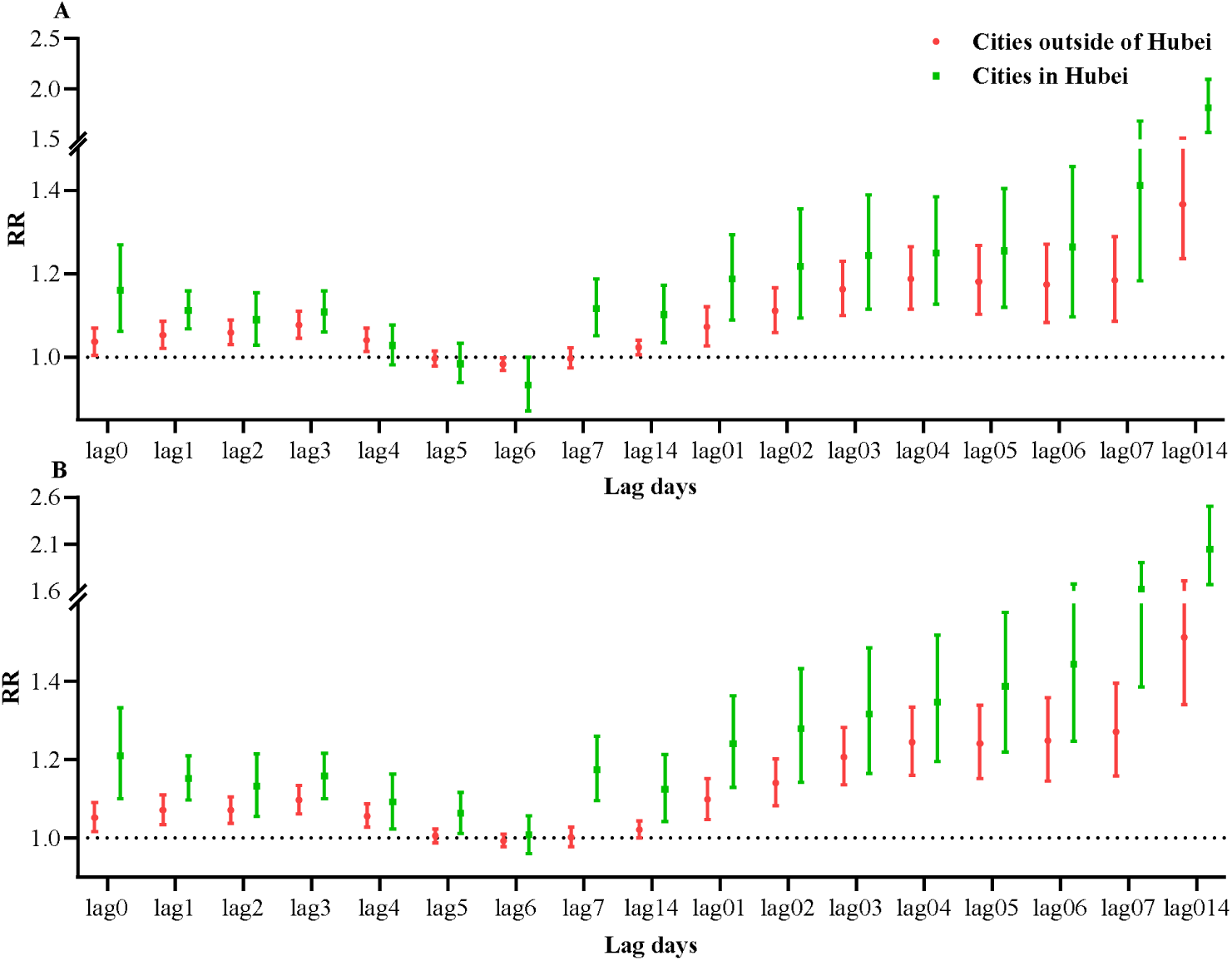
Associations between airborne PM pollution and confirmed cases stratified by regions in 72 cities of China during January 20 to March 02 2020 Note: (A) PM_10_; (B) PM_2.5_. The results were expressed as the relative risk (RR) and 95% confidence intervals (CIs) for the per 10 μg/m^3^ increase in PM_2.5_ and PM_10_ concentrations.

The fitted curves showed that the cities outside Hubei confirmed more COVID-19 cases around 30 μg/m^3^ for PM_2.5_ and 60 μg/m^3^ for PM_10_, similar trend were also found for the cities (12 cities) in Hubei cities. Association analysis indicated that both the cities inside Hubei and outside Hubei had the highest RR of COVID-19 in lag 014 for both PM_2.5_ and PM_10_. The stronger associations between COVID-19 and airborne PM were found for cities inside Hubei than outside Hubei.

## 4 Discussion

Aerosol has been recently confirmed as an transmission route for COVID-19 (van Doremalen et al. 2020), which may be modified by the level of air airborne PM pollution. To our knowledge, no studies have reported the associations between COVID-19 confirmed case counts and particulate pollution in cities across China. As of March 2^nd^, 2020, we collected a total of 24 939 confirmed cases from 72 cities in China and found positive relationships between PM_2.5_ and PM_10_ and COVID-19 cases after controlling meteorological factors and population migration.

As infection diseases, population migration might lead to the wide transmission among different regions, and our previous study found that the migration was positively related to the increase of daily increased confirmed COVID-19 case counts(van Doremalen et al. 2020). Meanwhile, because of evidently influence of meteorological factors in COVID-19 transmission (Wang et al. 2020b; Oliveiros et al. 2020) we chose to control meteorological factors and MSI in our models to clarify the associations between COVID-19 case counts and airborne PM pollution. After controlling these factors, we found both PM_2.5_ and PM_10_ were positively related to the COVID-19 confirmed cases, suggesting the airborne PM pollution might affect the COVID-19 transmission. This was similar to other studies focusing on the influenza and SARS. A study conducted in 47 Chinese cities has found that ambient PM_2.5_ may increase the risk of exposure to influenza in China especially during days with low temperature (Chen et al. 2017). Croft etc. have found that short-term increases in traffic and other combustion sources-related PM_2.5_ might contribute to the increased rates of influenza hospitalizations (Croft et al. 2020). Jaspers etc. reported that diesel exhaust increased influenza virus attached to respiratory epithelial cells within 2 h post-infection (Jaspers et al. 2005). Besides, long-range transport of influenza virus A was found during dust storms days with higher concentration of ambient influenza A virus (Chen et al. 2010). By conducting an ecologic study, Cui etc. found that case fatality rate of SARS in 5 regions increased with the increment of air pollution index (API) and those patients in regions with higher API suffered greater risks of death(Y et al. 2003).It is speculated that SARS-Cov-2 might also be transmitted by aerosols(van Doremalen N et al. 2020). Therefore, higher level of airborne particulate matters may increase the transmission of COVID-19.

PM pollution is a health hazard which could impair the immune function (Wei and Tang 2018). Research reported that airborne PM decreased the ability of pulmonary macrophages to effectively mount a defense against infection, which would last at least a week post-exposure via RelB activation (Migliaccio et al. 2013). Since pulmonary macrophages are very important in lung to phagocytize pathogene, the suppressed of that function would increase the invasive ability of SARS-Cov-2. In addition, airborne PM induces respiratory inflammation and affects the health of airway(Bateson and Schwartz 2004; Li et al. 2017). In particular, these severe inflammation in lung after exposure to PM_2.5_ were found to be mediated by angiotensin-converting enzyme 2 (ACE2), which showed significant increase in lung after PM_2.5_ exposure(L and TS 2020). Interestingly, it is reported that the receptor binding domain of the SARS-Cov-2 could be recognized by the extracellular peptidase domain of ACE2, which are predominantly expressed in a transient secretory cell type in subsegmental bronchial branches (Lukassen et al. 2020; Yan et al. 2020). Thus, the air PM may increase the possibility of SARS-Cov-2 lung invasion through ACE2 pathway. All these evidence together may explain why air airborne PM is positively related to increased risk of COVID-19.

To our knowledge, this is the first study to evaluate the association between air PM pollution and COVID-19 cases. Our findings indicate that airborne PM may be a key factor influencing the incidence of COVID-19 and more attention should be paid on the effect of air particulate pollution on COVID-19. Nevertheless, there are some potential limitations in this study. First, there are some other factors affecting the incidence of COVID-19, such as public health interventions, but we examined the impact of air pollution after controlling the population migration and meteorological factors. Second, there were changing COVID-19 case definitions at different stage of the epidemic, which may affect the confirmed counts. To reduce the bias caused by the changing definition, we included 72 cities with confirmed more than 50 cases in our analysis. In addition, the diagnosis of COVID-19 cases is much influenced by government screening standards, especially in Wuhan, so we didn’t include Wuhan in this study. Finally, the study was only conducted in China, while the COVID-19 is recognized as an emergent world pandemic, so our conclusions need future evaluation with global data. Despite of these limitations, our study provide some evidence from multiple cities across China and increased the knowledge over understanding the effect of PM pollution on COVID-19.

In conclusion, our results indicate that airborne PM likely increase the risk of having COVID-19 in China. However, the ecological fallacy and many uncontrolled confounding effects like different public health interventions may have biased our results. Further investigations including global data would be critical to study the association between COVID-19 and air pollution.

## Data Availability

The migration scale index(MSI) of each city was collected from Baidu migration map (https://qianxi.baidu.com/)

